# No-code machine learning in radiology: implementation and validation of a platform that allows clinicians to train their own models

**DOI:** 10.1101/2024.04.24.24306288

**Authors:** Daniel C. Elton, Giridhar Dasegowda, James Y. Sato, Emiliano G. Frias, Christopher P. Bridge, Artem B. Mamonov, Mark Walters, Martynas Ziemelis, Thomas J. Schultz, Bernardo C. Bizzo, Keith J. Dreyer, Mannudeep K. Kalra

## Abstract

Machine learning models can assist clinicians and researchers in many tasks within radiology such as diagnosis, triage, segmentation/measurement, and quality assurance. To better leverage machine learning we have developed a platform that allows users to label data and train models without requiring any programming knowledge. The technology stack consists of a TypeScript web application running on .NET for user interaction, Python, PyTorch, and MONAI for machine learning, DICOM WADO-RS to retrieve data from clinical systems, and Docker for model management. As a first trial of the system, researchers used it to train a model for clavicle fracture detection as part of an IRB-approved retrospective study. The researchers labeled 4,135 clavicle radiographs from 2,039 patients across 13 sites. The platform automatically split the data into training, validation, and test sets and trained a model until the validation loss plateaued. The system then returned a receiver operating characteristic curve, AUC, F1, and other metrics. The resulting model identifies clavicle fractures with 90% sensitivity, 87% specificity, and 88% accuracy with an AUC of 0.95. This model performance is equivalent to or better than similar models reported in the literature. More recently, our system was used to train a model to identify if ultrasound frames that contain personally identifiable information (PII). After validation, the model was used to help de-identify a large dataset that was to be used for research. This first-of-its-kind system streamlines model development and deployment and opens up an exciting new pathway for the use of AI within healthcare.

## 1. Introduction

The potential use-cases where artificial intelligence (AI) models can bring value to healthcare continues to grow. As of October 2023, the United States Food and Drug Administration (FDA) has has approved 692 AI algorithms.[1] However, this pace of AI adoption within healthcare has been slow.[2] One cause of this is that hospitals are cost strapped and still reeling from pandemic, and a tiny fraction of FDA-approved AI devices are covered by insurance.[3] A perhaps even bigger and more serious issue is that external validations of AI algorithms often show substantial drops in performance compared to what was originally reported in the FDA submission.[4, 5, 6] This degradation is understood to be caused by “distribution shift”.[7] Distribution shift may be caused by differences in scanner type, image acquisition protocols, or patient demographics. While multiple strategies and techniques have been proposed to address the distribution shift issue, the problem remains largely unsolved.[8]

Developing models in-house is an attractive proposition for hospitals as they can use their own datasets for training, which helps ameliorate the issue of distribution shift. The current push towards internalizing AI model development is coming in time of maturing ML platforms. Advances in graphical/tensor processing units (GPUs/TPUs) alongside improved cloud computing infrastructure have reduced the cost needed to train models. Mature software libraries such as the Medical Open Network for Artificial Intelligence (MONAI) and PyTorch make it easier than ever to train machine learning (ML) models on medical imaging data. Even so, code has to be written and choices have to be made regarding data preprocessing, data augmentation, neural network model architecture, model hyperparameter settings, batch size, stopping criteria, and validation criteria. Thus, the process of training ML models still requires the involvement of specialized personnel with programming expertise such as postdoctoral researchers, data scientists, or machine learning engineers. With industry salaries increasing relative to what healthcare systems can pay, finding staff to fill such roles is becoming more and more of a challenge.[2]

Several no-code platforms for training ML models exist from companies like Amazon, Apple, Calrifai, Google, and Microsoft.[9] Some of these platforms have been tested on publicly available medical imaging datasets.[9] However, to our knowledge none of these platforms support DICOM natively and they suffer from a lack of integration with hospital IT infrastructure. To use such services images must be pulled down, converted to image formats, and uploaded. Even after a model has been developed it then has to be packaged and integrated with clinical systems. It has been our observation that the entire process of training, validating, and deploying a ML model within healthcare is very time-consuming, involving coordination between many people and a complicated juggling of code and data across multiple systems.[3]. Lack of a standard process means that there are risks to patient data privacy and data security.

Research in autonomous machine learning (AutoML) has previously shown that many of the steps mentioned above can be automated.[10] No-code ML goes a step further than AutoML by providing a platform that empowers individual users with the ability to independently and iteratively train, validate, and deploy models without any programming required.[11] In this paper we present a novel no-code ML platform which allows physicians to create their own ML models for radiology. While our focus is on medical imaging (DICOM) data, the overall framework could be extended to other types of healthcare data such as electronic health records.

There are many reasons clinicians may want to develop models. Previously several of us trained an in-house model to identify improperly taken chest radiographs.[12] An early version of the ML software described below was used to train a classification model to classify mammography image view (CC vs MLO) and laterality (L vs R) to check for mistakes with the DICOM orientation tag (four classes total). A DenseNet121 model (8 M parameters) was trained on 1,789 mammography images downsampled to 128x128 pixels. The model achieved an average accuracy of 99.52% on a test set of 665 images. Based on that initial success with that codebase we then decided to develop it further into a no-code system so that users without coding experience could train similar models in the future.

The aforementioned model for mammography view classification is an example of a model for quality assurance. Models can also be implemented to assist in the areas of time-consuming, mundane tasks such as worklist prioritization/triage, the automation of time-consuming tasks such as the segmentation, and even to assist with diagnosis. Since our platform was developed in-house and the resulting models are only used internally, FDA approval is not required for such models to be used clinically. The lack of FDA approval for these models underscores the need for careful internal validation and oversight. Recently Panch et al. have argued that internal development of AI models is a promising pathway for AI use in healthcare.[13] As Panch et al. explain, hospitals have a strong obligation to set up their own validation and monitoring infrastructure regardless of whether models are developed internally or are FDA approved.

We tested our platform by providing the system to researchers at Massachusetts General Hospital. They ran a pilot study which used the system to train a clavicle fracture detection model for X-ray radiographs. Next we utilized the system to train a model that can identify ultrasound image frames that have baked-in personally identifiable information (PII). The resulting model was successfully used to remove images with PII in order to generate a fully deidentified dataset for research purposes.

## 2. System overview

Figure 1 shows an overview of our system. To ensure data privacy, the entire system is implemented on secured platforms within the hospital firewall. The data from the clinical vendor neutral archives (VNAs) may be de-identified and transferred to a research VNA. Alternatively images may be pulled directly from the clinical VNAs. During training and inference imaging data is not saved to disk. A load balancer insures that clinical operations are not effected by any data pulls.

**Figure 1:**
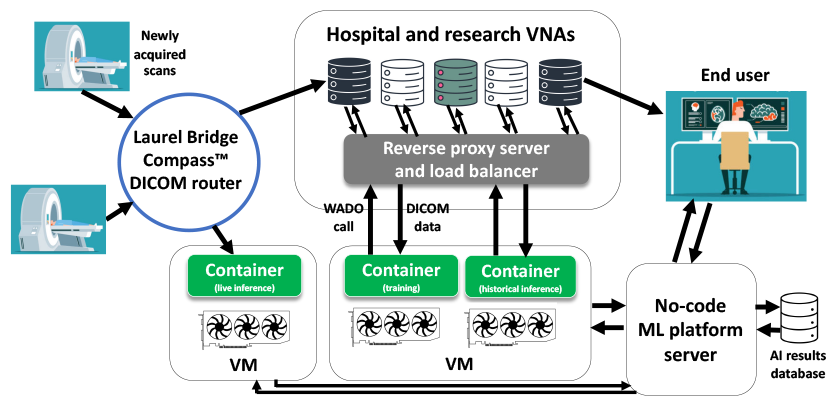
Platform overview.

The no-code platform graphical user interface (GUI) was written in TypeScript with a .NET core backend. Currently only classification models are supported. Users can use Nuance’s mPower software to search for studies to use and then import a list of studies into the system. This includes the ability to search by age and sex. A special GUI page for for data labeling was developed using the eUNITY viewing software to view images.

Users can also view the radiology text reports associated with each study. Users can provide labels at the both the study and image level. Once a training dataset has been created, then a training run can be started. A special interface was developed for training and model management. Training occurs in Docker containers which are run on dedicated virtual machines (VMs) with GPUs (NVIDIA A100s or V100s).

The Docker container contains code which uses the PyTorch ML framework and some functions from MONAI. For inputs, the container takes in a configuration file in Java Script Object Notation (JSON) format and a .csv file with labels and optional custom splitting (into training, validation, and test datasets). During training the container queries the hospital VNAs using Web Access to DICOM Objects (WADO) to retrieve images. The images are fed into the training on the fly. The trained model is saved in a local output directory alongside training logs and the original configuration .json file.

Training options can be modified by editing the JSON config file (in the future a GUI for this may be provided). While nearly all options in the system are configurable, the default settings were carefully chosen so they should be suitable for most applications.

The default model is a pretrained DenseNet201 architecture (20.1 M parameters), but the system supports many models which are available from MONAI, such as EfficientNet models, which use less compute.[27, 28] The user must specify the number of classes and whether they are doing exclusive classification, non-exclusive classification, or regression.

By default both dropconnect[29] and dropout are used with a conservative rate of 0.2 and the following types of data augmentation are implemented - random rotations (−10 to +10 degrees), random zooms/crops (0.9 - 1.25x), random flipping, and random elastic deformations. The amount of elastic deformation is kept small as large deformations may not be suitable for all applications. Several options for data normalization are provided (clipping, rescaling, etc). Models are trained using the Adamax optimizer algorithm,[30] with weighted random sampling to improve performance in the case of unbalanced training labels. The default batch size is 6 and the default learning rate is set low at 0.0005 to avoid training instabilities. We use the well-known “reduce on plateau” learning rate schedule, which reduces the learning rate by a factor of 0.5 when the validation loss plateaus for 5 steps.

A key challenge in no-code ML is determining the criteria for stopping training. If the model is trained too long it will overfit, whereas if training is stopped too early the model’s accuracy may not have reached the best possible value. Since the validation loss can be very noisy we smooth the validation loss using a moving average of width 5 iterations. We stop the training when the smoothed validation loss no longer decreases for 1500 iterations. The maximum number of epochs is set to 400.

The system contains several options for train-validation-test splitting. The default is to split the data by *PatientID*. The splitting can also be done by site (hospital) which is stored in the *IssuerOfPatientID* DICOM tag. Thus, data sources in the test set can be different from those in the training set, providing a form of “external” validation. Once a model has been trained, it is run on the test set. The system returns validation statistics, ROC curves, and a confusion matrix (see fig. 3.3. The Youden index is used to determine the optimal threshold for classification during inference.[31]

**Figure 2:**
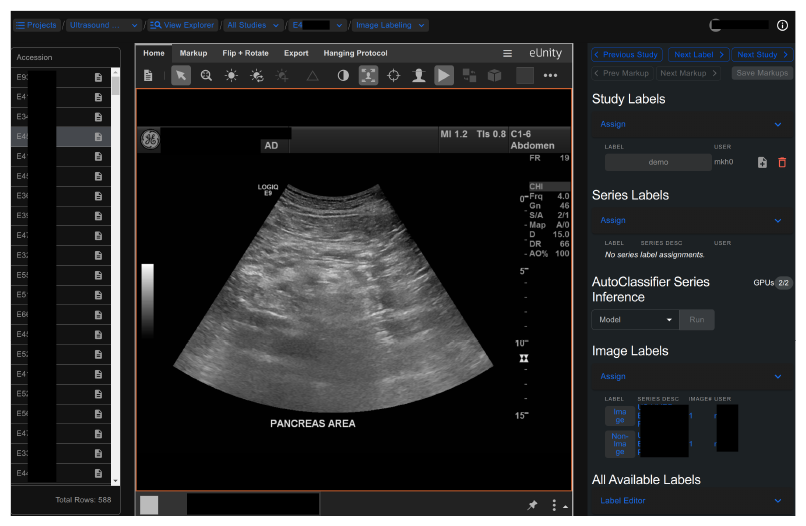
The interface for data labeling.

**Figure 3:**
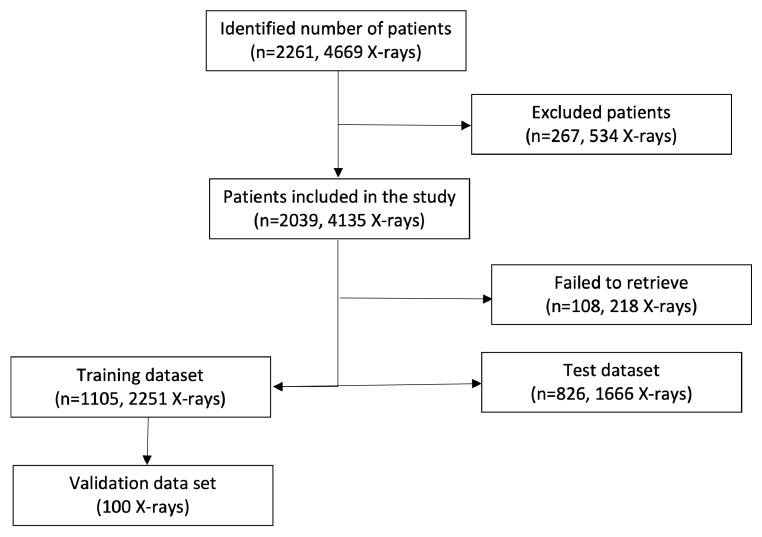
Flowchart of the inclusion and exclusion criteria for the training, validation, and test datasets for the pilot study.

After a model has been trained and tested, new containers for inference may be spun up. Inference can be done on historical data listed in a .csv or live-routing may be configured. During live-routing the newly acquired scans that match set critiera are routed to the container. Currently live routing must be manually configured using a custom C# script implemented in our Laurel Bridge Compass™ DI-COM router. Any potential clinical use of models developed with our framework requires approval by our AI governance board and extensive period of validation with live data.

## 3. Pilot user study

### 3.1. Study design

Our institution review board (IRB) waived the written consent requirement for our retrospective, Health Insurance Portability and Accountability Act (HIPAA) compliant pilot study. The study details provided below are presented in conformance with the Checklist for Artificial Intelligence in Medical Imaging (CLAIM).[32]

The study dataset was comprised of clavicle radiographs from 2,039 adult patients (age > 18 years) sourced from 13 sites within our network including 10 hospitals, one urgent care center, and two quaternary care centers. To identify eligible radiographs for our study we used Nuance mPower Clinical Analytics Search (Microsoft Inc.), a cloud-based, commercial radiology reports search engine that integrates radiology reports data from the sites included in our study. The search key terms for identifying consecutive radiology reports and radiographs with and without clavicle fractures were “acute fracture” OR “no fracture” OR “displaced fracture” AND “clavicle X-ray”. The search was limited to clavicle radiographs performed between January 2016 – December 2022. Both right and left clavicle radiographs were included in the study. A post-doctoral radiology research fellow (2 years of experience) reviewed all radiology reports and radiographs to exclude clavicle radiographs with incomplete anatomic coverage of clavicles, metal-related artifact, prosthesis, or evidence of open reduction with internal fixation (*N* = 267). Non-clavicle radiographs (such as shoulder and chest radiographs) with or without clavicle fractures were not included in the study. A flowchart of the inclusion and exclusion with training and test data has been represented in fig. 1.

We exported radiology reports of the eligible radiographs from the radiology report search engine with the following data elements: radiology findings text, radiology impression text, date of examination, name of the radiographic procedure, site of radiographic acquisition, as well as patients’ age and gender. We reviewed the radiology reports and recorded the details of the presence of fracture to establish the ground truth. For this initial pilot study the data labeling was not done with the platform’s GUI. Instead, the data labels were uploaded via a .csv file into the system. To avoid selection bias, all consecutive clavicle radiographs were included regardless of patients’ age, race, and sex as well as radiographic equipment and site of acquisition. Although a power analysis was not performed to determine adequate sample size for our test set, our sample size was larger than most prior publications in this domain. Clavicle radiographs from three sites were marked for inclusion in an external test dataset. The remaining radiographs were used for training and validation.

### 3.2. Dataset and model

A total of 2,151 clavicle radiographs were used in the training dataset, and 100 radiographs from were automatically split to comprise the validation dataset. The default settings described in section 2 were used. All radiographs were resampled to a size of 512x512 and normalized to have a mean of zero and standard deviation of one. After model training and testing the platform returned statistics for the performance of the AI model. The mean age (± standard deviation) of 2039 adult patients included in our study was 52 years. There were 1022 female patients and 1017 males. Site-wise distribution of patients was Site 1 (N=621 patients), Site 2 (N=651), Site 3 (N=166), Site 4 (N=40), Site 5 (N=223), Site 6 (N=167), Site 7 (N=43), Site 8 (N=11), Site 9 (N=6), Site 10 (N=36), Site 11 (N=37), Site 12 (N=7), and Site 13 (N=31). Of the 2039 x-rays, there were 1225 radiographs of the right clavicle and 814 left clavicle radiographs. Most patients were either outpatients (N=1066) or in the emergency department (N=828), with only 145 inpatients. Radiographs of 108 patients who could not be automatically deidentified were excluded. 1,666 radiographs were used for model testing (772 radiographs with clavicle fracture and 894 radiographs without clavicle fracture) from two sites that did not contribute to the training datasets. The test set included 826 patients total.

### 3.3. Results

The model trained until it achieved 89% accuracy on the validation set. On the test set the model classified individual X-ray images with clavicle fractures with 90% sensitivity, 87% specificity, 88% accuracy, 0.86 F1, and an area under the reciever operating characteristic curve (AUC) of 0.95. The model had 91% sensitivity, 94% specificity, 93% accuracy, 0.91 F1, and an AUC of 0.97 (95% CI 0.96-0.98) when classifying at the patient level (since some patients had multiple images in their study, we average the softmax predictions across the images for each patient and use that as a ‘patient level’ classification). Receiver operating characteristic (ROC) curves are shown in fig. 3. Comparison with some previous models for fracture detection is provided in table 1. Additional statistical analyses were performed with SPSS. There was no significant difference in model performance in male or female patients and among patients in different locations at the time of their radiography (*p* > 0.05). For analysis, AI outputs were classified as true positive, true negative, false positive, and false negative. False positive findings were noted in X-rays when the clavicle had degenerative changes, skin folds, artifacts, and foreign bodies such as post catheter overlying the clavicle. False negative findings were present in both displaced and non-displaced clavicle fractures. Examples of true positive, true negative, false positive, and false negative outputs are shown in fig. 3.2.

**Table 1.**
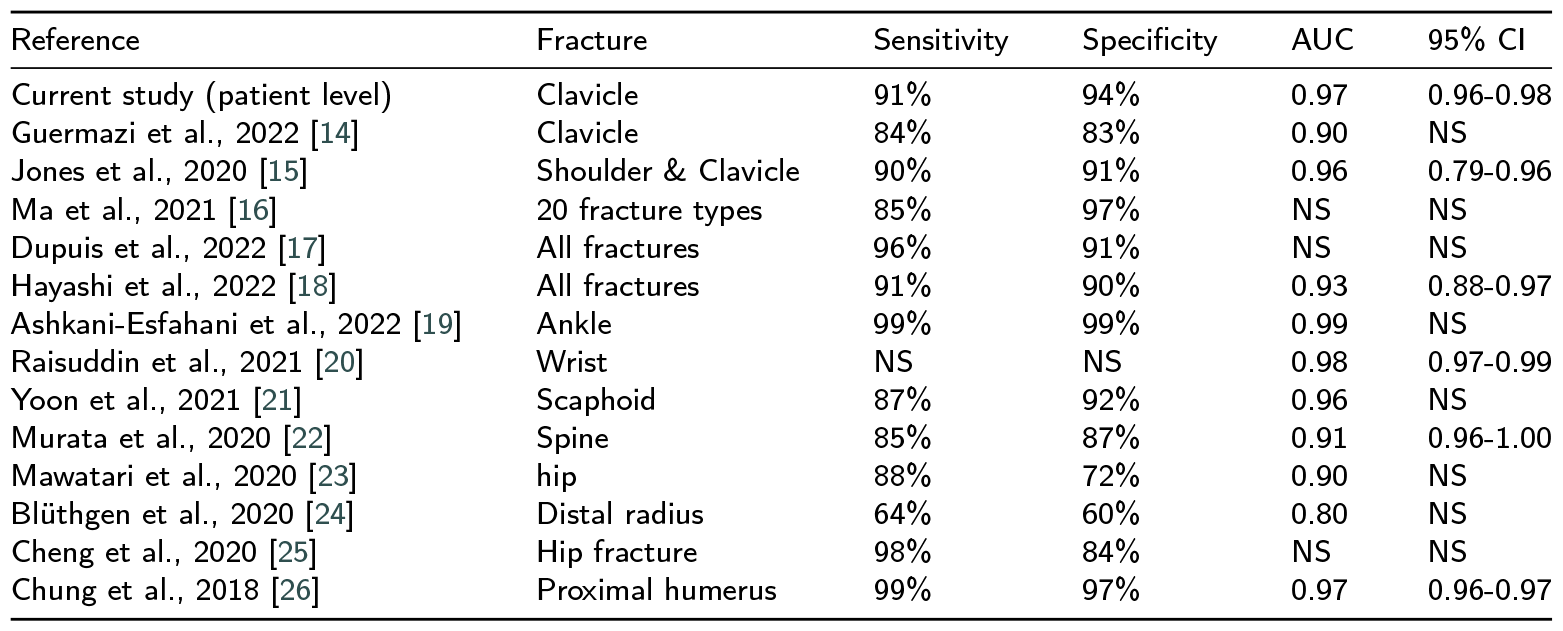
Summary of some previously published fracture detection models. (NS = Not specified)

## 4. First use-case: identifying PII in ultrasound images

The first use-case of the fully developed system was to train a model to classify ultrasound images as to whether they were “normal” images, “non-images”, or images with burned in PII. Examples of these image types are shown in figure 4. Ultrasound images with burned in PII are rare but must be removed when creating fully de-identified datatsets for research purposes. Previously this time-consuming process was done manually.

**Figure 4:**
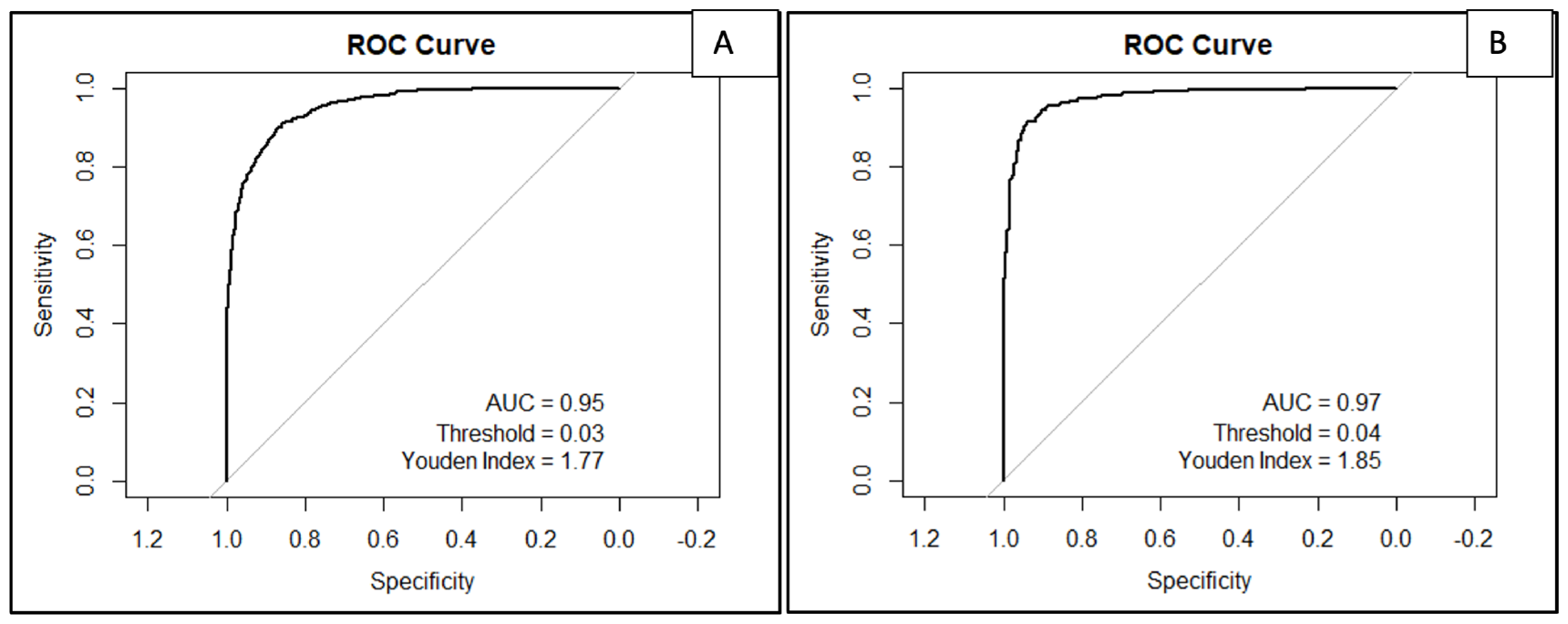
ROC curves and AUCs for X-ray level (A) and patient level (B) clavicle fracture detection.

**Figure 5:**
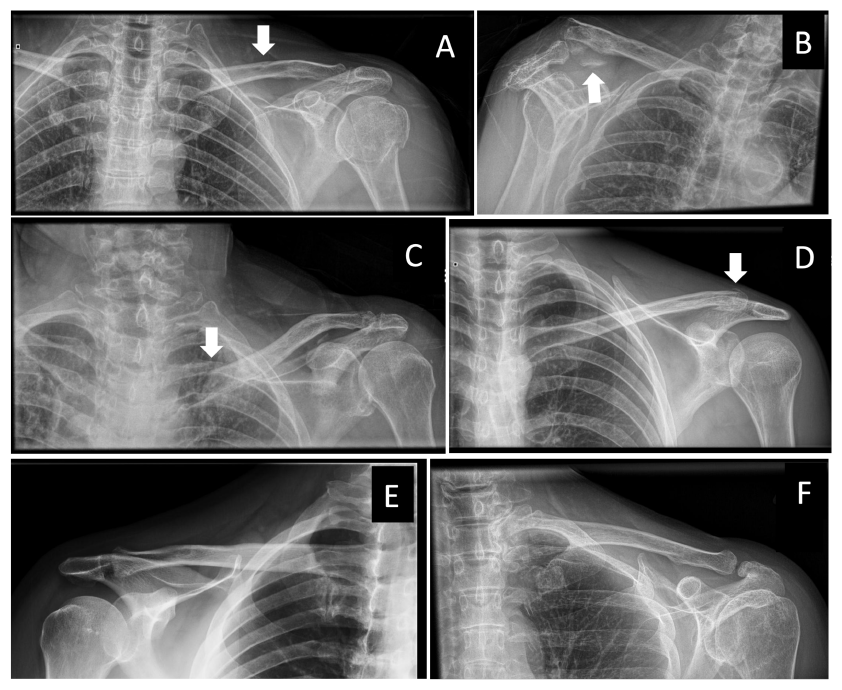
Model performance examples on frontal projection radiographs of clavicles with AI-detected (true positive: A, B), AI-missed (false negative: C, D), and AI-false positive (E, F) clavicle fractures

**Figure 6:**
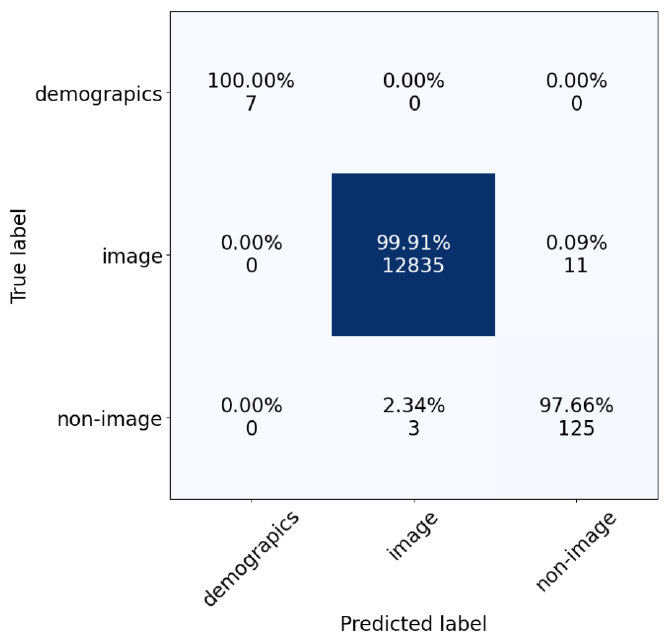
Confusion matrix on a test set of ultrasound images.

**Figure 7:**
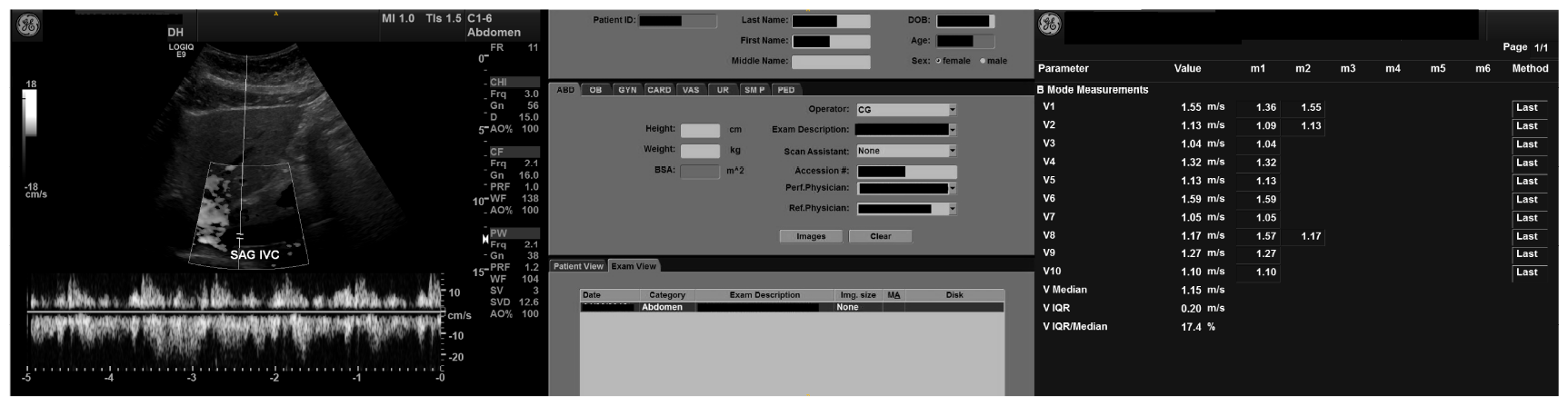
Examples of a standard ultrasound image frame (left), an ultrasound image which is actually a screenshot with PII (middle), and an ultrasound image that is a “non-image” settings page. (PII has been redacted in black).

**Figure 8:**
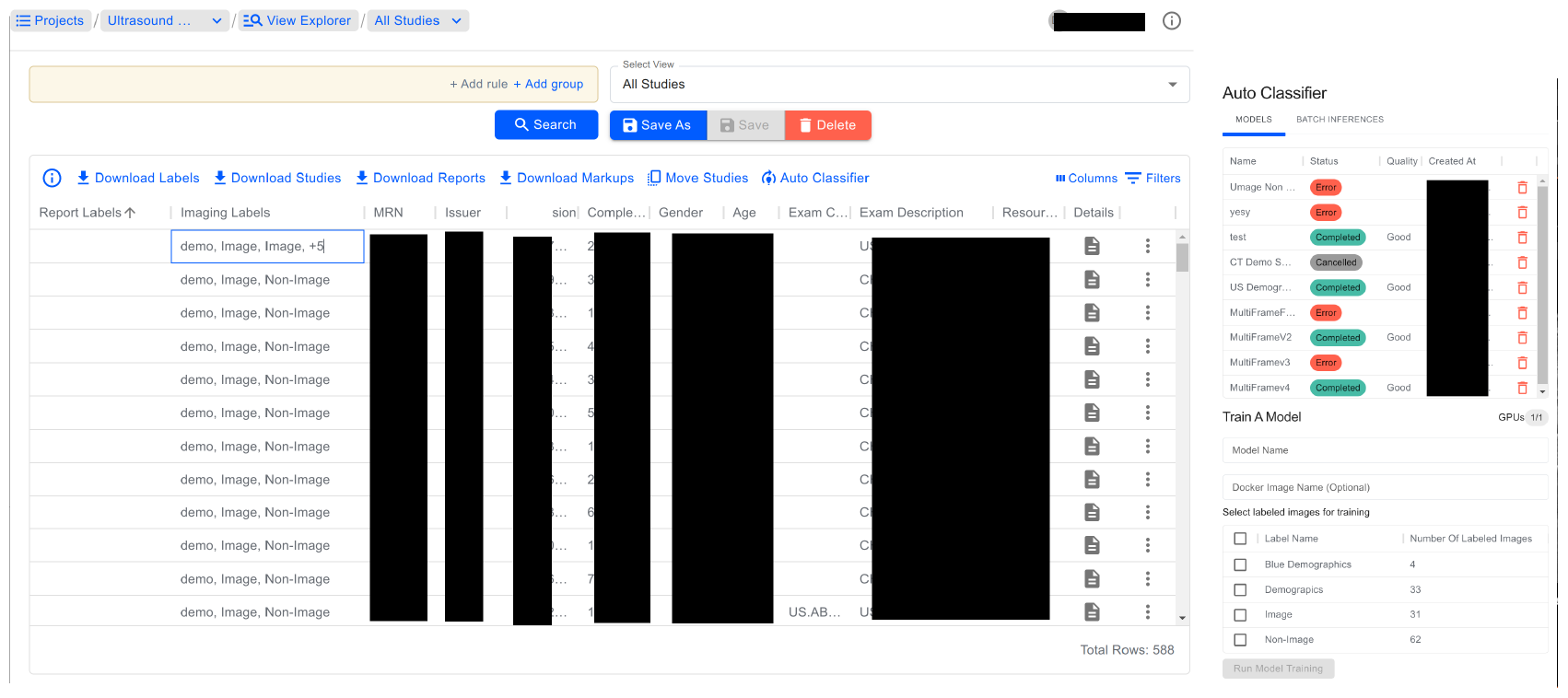
Screenshots of the data list (left) and ML model management pane (right).

To properly test the system, the model development was done by a program manager with no technical coding knowledge. Using the system a training set had 421 images (including multiframe) were labeled. There were 62 “non-image” images, and 37 images with PII. While these numbers are small, the classification task is relatively easy due to the large differences between the images types. The “normal” images were DICOM MultiFrame images (sometimes called “Enhanced DICOM”). The multiframe images contained around 20-30 frames each. Since the system trains on all frames, the total number of “normal” images was around 9,500.

The test set consisted of 12,981 ultrasound instances, many of which were multiframe. Manually labeling the test set with our GUI was found to be time consuming, so it was done by downloading all of the data and viewing image thumbnails. This enabled the dataset to be labeled within hours rather than days. The confusion matrix is shown in fig. 3.3. The overall accuracy was 0.9989. While the number of demographics pages in the test set is small, these initial results were encouraging enough that the model was used to help de-identify a separate dataset for research purposes.

## 5. Discussion

Our study demonstrates that an internally developed no-code ML platform give non-coding clinicians and personnel the ability to develop AI classification models in a fast and efficient manner. The performance of the clavicle fracture classification model trained with our system is similar or better than models reported in the literature (see table 1.[33, 34] For example, Guermazi et al. reported their AI model for identifying shoulder and clavicle fractures had a performance of 84% sensitivity, 83% specificity, and 0.90 AUC (95% CI 0.79-0.96).[14] Jones et al. reported on a deep learning system for identifying clavicle fractures, using an ensemble of 10 convolutional networks, which obtained 90% sensitivity, 91% specificity, and an AUC of 0.96.[15]

The clavicle detection model pilot study has some notable limitations. First, there was an asymmetric distribution of radiographs across different institutions and between the radiographs with and without clavicle fractures. Second, all clavicle radiographs belonged to a common healthcare system in the same geographic location in the Northeast part of the United States. Third, we did not assess variations in the model performance across patients’ size, racial, or ethnic groups. Fourth, as stated above, the successful creation of a classification model for clavicle fractures does not imply that the same no-code ML platform will be successful at building sophisticated models or those for cross-sectional imaging modalities. Furthermore, model performance on different radiography techniques (i.e. computerized versus digital radiography) was not assessed; however, considering variations in equipment across the 14 hospitals, the model was trained and tested on radiographs from several different vendors. Likewise, due to the exclusion of radiographs with incomplete anatomic coverage, artifacts, and prior open reduction and internal fixation, we cannot comment on the model performance on such radiographs. Finally, we also did not assess the model’s performance in non-displaced versus displaced fractures and for patients with chronic or non-healed clavicle fractures. Despite these limitations, we believe this model could improve the diagnostic accuracy of fracture detection, especially in clavicle fractures without displacement that often pose a challenge to interpreting physicians.

There are several avenues open to improving this system. One thing that still needs to be implemented is better support for cross validation. Currently cross-validation must be done by providing a “fold” parameter in the config file. Then the training runs for each fold have to be initiated manually, and the metrics for each fold have to be averaged manually as well. This process could be automated on the web platform side. Next, the system could be expanded to work with 3D modalities such as CT or MRI. Since we have already implemented training on multiframe images, this should be easy to implement. A more substantial upgrade would be to extend the system beyond classification to enable segmentation and bounding box detection models to be created. We have already extensively tested the Redbrick AI (www.redbrickai.com) data labeling platform and integrated it with our VNA. Redbrick AI provides an application programming interface (API) which allows cohorts to be sent the platform. On the platform users can create segmentation, polygon, and bounding box labels which can then be retrieved via API. The image labeling platform could also be updated to make labeling faster and easier in some situations, for instance by implementing thumbnail views. Finally, in the future this system could be further developed to allow users to fine-tune multimodal transformer foundation models similar to GPT-4.

In conclusion, our no-code ML platform simplifies and expedites the development of AI models for medical imaging. It is our goal to make this system available to scientists, physicians, and engineers within our healthcare system so they can train models to assist in their everyday work.

## Data Availability

All data produced in the present work are contained in the manuscript. Supporting spreadsheets are available upon reasonable request. The imaging data used for training will not be available.

